# Lower pre-treatment TMS-evoked cortical reactivity and alpha-band oscillatory dynamics predict efficacy of primary motor cortex neuromodulation for chronic pain

**DOI:** 10.1101/2025.10.11.25337793

**Authors:** Enrico De Martino, Margit Midtgaard Bach, Anne Jakobsen, Bruno Nascimento Couto, Nahian S. Chowdhury, Stian Ingemann-Molden, Adenauer Casali, Thomas Graven-Nielsen, Daniel Ciampi de Andrade

**Affiliations:** Center for Neuroplasticity and Pain (CNAP), Department of Health Science and Technology, Faculty of Medicine, Aalborg University, Aalborg, Denmark; University of New South Wales, Sydney, New South Wales, Australia; Pain IMPACT, Neuroscience Research Australia, Sydney, Australia; Institute of Science and Technology, Federal University of São Paulo, São Paulo, Brazil

**Keywords:** Repetitive transcranial magnetic stimulation (rTMS), neuromodulation, Chronic pain, TMS-EEG, Cortical excitability, Biomarkers of treatment response, brain oscillations, neuropathic pain

## Abstract

Repetitive transcranial magnetic stimulation (rTMS) to the primary motor cortex (M1) provides significant pain relief in ∼45% of chronic pain patients. Identifying biomarkers that predict treatment response before starting rTMS is essential for guiding clinical decision-making. Here, we used TMS combined with electroencephalography (TMS-EEG) to assess pre-treatment cortical function in 43 patients with chronic pain before receiving 12 sessions of therapeutic 10 Hz rTMS to M1 over eight weeks as a secondary analysis from a trial comparing effects of rTMS in different cortical targets. Responders were defined as individuals reporting a ≥30% reduction in pain intensity on a visual analogue scale at week 8. Pre-therapy TMS-evoked cortical reactivity was quantified at M1 using global mean field power (GMFP), and local mean field power (LMFP). Oscillatory dynamics were measures by event-related spectral perturbation (ERSP), and intertrial coherence (ITC) in alpha (8–12 Hz), low-beta (13–20 Hz), and high-beta (21–30 Hz) bands. Compared with non-responders, responders (n=20; 47%) showed lower GMFP, LMFP, alpha-band ERSP, and ITC at the stimulation site (all p<0.05). These low measures correlated with greater reductions in pain intensity (p<0.05). Exploratory supervised machine-learning analysis using three TMS-EEG features (GMFP, alpha-band ERSP, alpha-band ITC) predicted responder status with acceptable performance (ROC-AUC = 0.70, PR-AUC = 0.76). These findings suggest that lower pre-treatment TMS-evoked cortical reactivity and alpha-band oscillatory dynamics may identify patients more likely to benefit from rTMS. Prospective clinical trials should test pre-therapy reactivity and connectivity metrics to select patients more likely to benefit from therapy.

## Introduction

High-frequency (10 Hz) repetitive transcranial magnetic stimulation (rTMS) of the primary motor cortex (M1) is a non-invasive neuromodulation technique that induces analgesic effects in patients with chronic pain [24]. RTMS repetitively depolarizing axons [43], inducing activity-dependent plasticity in corticothalamic networks [22], and increases phase-, and power-based metrics of functional connectivity that outlast the stimulation session [29]. International guidelines currently recommend rTMS for the management of different chronic pain conditions, such as neuropathic pain [45], migraine [34], and fibromyalgia [44]. However, clinical trials have demonstrated high inter-individual response variability in chronic pain patients receiving M1 rTMS treatment, with only approximately 45% showing a pain reduction of more than 30% [1,36]. Determining the likelihood of treatment response at the individual level is highly relevant in clinical practice, as it may help direct therapy to those who are more likely to benefit from it [10]. A recent feasibility pilot study combining transcranial magnetic stimulation with electroencephalography (TMS-EEG), a neurophysiological approach for assessing cortical function [48], showed that pre-treatment global phase-based connectivity may detect responders to M1 rTMS therapy for pain relief. Although innovative, this feasibility report explored only a single, global phase-based connectivity measure and opened the venue for a more comprehensive assessment of markers of response to therapy based on readouts of pre-treatment cortical oscillatory dynamics [39]. TMS-EEG enables the assessment of many aspects of cortical physiology, including TMS-evoked global and local reactivity and TMS-evoked oscillatory dynamics, through measures such as global mean field power (GMFP), local mean field power (LMFP), event-related spectral perturbation (ERSP, a measure of changes in power across frequencies in response to stimuli), and intertrial coherence (ITC, a measure of local phase reset and consistency across trials) [48]. These metrics provide a comprehensive assessment of functional connectivity across global and local cortical circuits after the delivery of a probing TMS pulse, providing insights into cortical excitability, and power/phase-based connectivity in specific spectral bands. Previous studies have shown that experimental acute pain reduced both local M1 reactivity (LMFP measured peri-cortical target) and global reactivity (GMFP) compared to non-painful stimuli [28]. Additionally, reductions in amplitude-based (ERSP) and phase-based (ITC) cortical oscillatory dynamics were also shown to be reduced during acute pain in the alpha band following M1 stimulation and in the low beta band following left dorsolateral prefrontal cortex (DLPFC) stimulation [27], suggesting a less synchronized and more variable cortical response to TMS probing of both M1 and DLPFC during short-lasting experimental pain. Notably, decreases in TMS-evoked oscillatory dynamics did not occur uniformly across participants: approximately 60% of healthy individuals exhibited predominant reductions in local cortical function in either the DLPFC or M1, but rarely at both sites concurrently [28]. Additionally, individual trait sensitivity to pain was significantly related to the magnitude of M1-evoked oscillatory dynamics during experimental pain, with lower reductions in phase-based connectivity correlating with higher individual pain sensitivity [27]. These data consistently suggest that even in healthy individuals under experimental pain conditions, personal characteristics may influence how pain affects cortical function [28] in an individual-specific manner. Although those findings concern experimental pain in healthy participants, they suggest trait-like differences in state-dependent M1 responsiveness that could be generalizable to patient populations. It would therefore be possible to hypothesize that individuals with different profiles of TMS-EEG M1 responses respond differently to modulatory therapeutic interventions aiming at this brain area. Based on homeostatic plasticity principles, which propose that cortical activity is regulated via compensatory adjustments whenever it deviates from the optimal range [2], we hypothesized that chronic pain patients with low cortical reactivity and oscillatory dynamics upon probing M1 before treatment would be more likely to benefit from therapeutic M1 rTMS compared to those without these features. To test this hypothesis, we compared TMS-evoked cortical reactivity, as well as oscillatory dynamics, measured before therapy to treatment outcomes in chronic pain patients receiving M1 rTMS Moreover, we provided single-patient classification perspective for each TMS-EEG metric based on reference data from 125 healthy pain-free individuals.

## Methods

### Patients

This study is a secondary exploratory analysis from a larger trial that assessed the use of pre-treatment TMS-EEG to select the cortical target for rTMS in each patient. In this double blind, three-arm randomized clinical trial (clinical trial registration number NCT06395649) [30] M1 was used as a target in the active control comparator arm, representing the current best neuromodulatory treatment. Among 90 patients with chronic pain, 43 patients received rTMS treatment targeting M1 and were analyzed in the present study. Eligible participants were males and females aged 18 to 80 years who had experienced pain persisting for at least 6 months. Patients with both primary and secondary pains, covering all mechanistic descriptors of pain (neuropathic, nociplastic, and nociceptive) with at least moderate pain (pain intensity higher than 3/10 on a 0–10 numerical rating scale; NRS) were allowed in the trial. Exclusion criteria included diagnosed psychiatric conditions, substance abuse, unstable medical conditions, contraindications to rTMS, such as severe head trauma, prior neurosurgery, current or past epilepsy, intracranial hypertension, or implanted ferromagnetic devices. Additional reasons for exclusion included inability to complete study follow-up, current pregnancy, or breastfeeding. The study was approved by the local Ethics Committee (N-20230076) and conducted in accordance with the Helsinki Declaration. Written informed consent was obtained before the start of the study.

### Study design

After recruitment and before rTMS, patients participated in a TMS-EEG session probing M1 with single-pulse TMS under EEG recording. rTMS sessions began the week after the TMS-EEG assessment and lasted 8 weeks. The protocol included five sessions over five consecutive weekdays (induction phase), followed by a maintenance phase with one session per week for seven weeks, totaling 12 rTMS sessions. At the end of the study, participants were divided into Responders or Non-responders based on their level of pain reduction and used for the current study. Responders were defined based on a predefined primary outcome by reduction of more than 30% in mean pain intensity, calculated by comparing scores on the last day of the maintenance phase (week 8) with the average pain intensity during the week preceding the TMS-EEG assessment. Pain intensity was assessed using a visual analogue scale (VAS), which allows participants to rate their pain on a continuous scale ranging from 0 to 100, where ‘0’ indicates ‘no pain’ and ‘100’ indicates ‘the worst pain imaginable.’

### Clinical trial outcome

The primary outcome measure was the reduction in pain intensity, evaluated after each rTMS session using the VAS [4], referring to the patient’s pain at that moment in time. Furthermore, at the beginning and the end of the study, additional questionnaires were completed by the patients, including average pain over the last 7 days, pain in the last 24 hours, a body chart, the short-form brief pain inventory (BPI), the hospital anxiety and depression scale (HADS), and quality of life (EQ-5D), the number and type of medications, sleep quality, fatigue, and patient global impression of change (PGIC).

From the BPI, two indices were extracted: pain severity, calculated as the average of four pain intensity items (worst, least, average, and current pain), and pain interference, calculated as the average of seven items assessing interference with general activity, mood, walking ability, normal work, relations with others, sleep, and enjoyment of life. The number of painful body regions was calculated based on participants’ reported body maps to quantify the spatial distribution of pain [47]. The HADS is a 14-item self-report questionnaire designed to assess symptoms of anxiety and depression, with higher scores indicating greater symptom severity [49]. The EQ-5D is a measure of health-related quality of life, assessing five dimensions: mobility, self-care, usual activities, pain/discomfort, and anxiety/depression. Each domain is rated by the participant based on perceived problems using a VAS [12]. Sleep quality and fatigue were also assessed using VAS. For sleep, participants rated the question ‘How did you sleep last night?’ on a scale from 0 (bad sleep) to 100 (good sleep). For fatigue, they rated the question ‘How tired have you been in the last 24 hours?’ on a scale from 0 (no tiredness) to 100 (worst imaginable tiredness).

The PGIC is a 7-point Likert-type scale used to assess a participant’s subjective evaluation of overall improvement following treatment. Responses range from “Very much worse” to “Very much improved”, capturing the individual’s perceived change in health status during the trial [19].

### TMS-EEG assessment

Two-hundred single-pulse TMS was delivered using a biphasic stimulator (MagPro R30, MagVenture A/S, Farum, Denmark) and a figure-of-eight coil (Cool-B35, MagVenture A/S, Farum, Denmark) [29]. TMS was applied to the hemisphere contralateral to the side of the most intense pain. If patients did not report a predominant site, stimulation was delivered over the left hemisphere.

To record TMS-evoked potentials (TEPs), a TMS-compatible passive electrode cap with 63 electrodes (10-20 system; g.tec-medical engineering GmbH, Schiedlberg, Austria) was used, with the Cz electrode positioned at the vertex. The ground electrode was placed on the right zygoma, and a reference electrode was located on the right mastoid [28]. Electrode impedance was monitored and kept below 5 kΩ. Raw signals were sampled at 4800 Hz using a TMS-compatible EEG amplifier (g.HIamp, g.tec-medical engineering GmbH, Schiedlberg, Austria). To minimize auditory responses from the click generated by the TMS coil from interfering with the TEPs, a TMS-click sound masking toolbox (TAAC, [40]) was used, and participants wore noise-canceling in-ear headphones (ER3C, Etymotic, 50 Ohm, Illinois, USA). To reduce somatosensory sensations caused by the TMS coil, two net caps (GVB-geliMED GmbH, Ginsterweg Bad Segeberg, Germany) with a plastic wrap were applied to the EEG [28].

A navigated brain stimulation software (InVesalius Navigator, 3.1) was used to calibrate the participant’s head and the TMS coil position with the aid of an optical (infrared) motion capture camera (Polaris Vicra, NDI, Ontario, Canada) [46]. The M1 target was identified by locating the hand muscle representation in the left or right hemisphere at the largest observable hand movement. The rMT was determined as the TMS intensity needed to evoke a visible hand movement. An intensity of 90% of the rMT was used for the TEPs to prevent sensory feedback contamination [13]. Furthermore, to ensure a detectable TEP, a real-time visualization tool (rt-TEP) was used [6]. This interface enabled adjustments to the TMS coil orientation and intensity across participants to minimize unwanted artifacts and ensure a minimum early peak-to-peak amplitude of 6 μV, as measured in the average of 30 trials at the EEG electrode closest to the M1 target. The InVesalius Navigator system and rt-TEP tool were used throughout the study to monitor the TMS coil position (keeping it within 3 mm of the cortical targets), with the interstimulus interval randomly jittered between 2000 and 2400 ms to prevent substantial reorganization or plasticity processes from affecting longitudinal TMS-EEG measurements [7]. A total of 200 single-pulse TMS stimulations were administered to each patient. During TMS stimulation, participants sat comfortably on an ergonomic chair with their eyes open, focused on a fixation point on the wall.

### TMS-EEG analysis

The pre-processing was performed using Python (Python Software Foundation). EEG data were segmented into epochs of ±800 ms around the TMS trigger. The signal between 0 and 15 ms containing the TMS artifact was replaced by temporally mirrored data from the pre-stimulus period [30]. Epochs and channels containing decay artifacts, 50 Hz line noise, eye blinks, eye movements, or facial and neck muscle activity were visually inspected and manually rejected, ensuring the highest possible signal quality while retaining a minimum of 150 trials. A single independent component analysis (fast ICA) was used to remove residual artifacts [25]. Finally, TEPs were band-pass filtered between 1 and 80 Hz using a third-order Butterworth filter, down-sampled to 1200 Hz, EEG channels re-referenced to the average reference, and any bad channels were interpolated using spherical splines [11].

Approximately half of the patients received stimulation over the left hemisphere (due to right-sided pain or bilateral pain), and the other half received stimulation over the right hemisphere (due to left-sided pain). To enable consistent group-level comparisons, data were reorganized according to stimulation laterality. Specifically, results were analysed relative to the stimulated hemisphere, regardless of whether stimulation was applied to the left or right hemisphere.

A predefined time window of 20–120 ms post-TMS was selected for extracting TMS-EEG outcomes of TMS-evoked cortical reactivity and TMS-evoked oscillatory dynamics. This early interval has been adopted in prior TMS-EEG studies because it captures the most specific cortical responses to TMS [27,29], while minimizing contamination from remaining somatosensory and auditory evoked potentials, which are most pronounced at 180-250 ms [37].

### TMS-EEG outcomes

To assess global cortical reactivity to TMS, GMFP was calculated as the root-mean-squared value of the TEP across all electrodes following TMS stimulation [28]. This measure reflects the overall strength of TEPs across the entire cortex, providing insight into widespread cortical activity following stimulation [21]. Furthermore, to assess local cortical reactivity to TMS, LMFP was calculated for the local cluster of electrodes, thereby focusing the analysis on the direct effects of TMS [20]. Specifically, the central clusters consisted of C1, C3, CP3, and CP1 on the left hemisphere, and C2, C4, CP2, and CP4 on the right hemisphere. To assess TMS-evoked oscillatory dynamics in these clusters in the stimulated hemisphere, time-frequency maps were computed in the 8–45 Hz range using Morlet wavelets with 3.5 cycles [27]. The analysis focused on three frequency bands: alpha (8–13 Hz), low beta (14–20 Hz), and high beta (21–30 Hz) based on previous study results [27,29]. Two main parameters were extracted: ERSP and ITC. ERSP was used to quantify changes in power across time and frequency over trials. It was computed as the average spectral power ratio of individual EEG trials relative to a pre-stimulus baseline (−600 to −50 ms). This measure captures how TMS-evoked oscillatory power evolves across specific frequency bands. Statistical significance was determined by comparing ERSP values to baseline using bootstrapping (500 permutations, two-sided test, p < 0.05, with FDR correction for multiple comparisons). Significant ERSP values were then averaged across time points and electrodes to characterize frequency-specific power modulation evoked by TMS [27,29]. ITC was extracted to evaluate phase-reset of cortical oscillatory activity across trials, serving as a marker of phase synchronization. ITC was calculated by normalizing the complex-valued time-frequency data from each trial, averaging across trials, and then computing the absolute value. Higher ITC values (approaching 1) reflect stronger phase-locking to the TMS pulse and indicate greater neural synchrony across trials. Significance was assessed using bootstrapping against the baseline (500 permutations, one-sided test, p < 0.05, FDR-corrected), and significant ITC values were averaged across electrodes, time windows, and frequency bins [27,29].

### Reference data

For the same outcomes, a control group of 125 healthy pain-free participants (age range: 20– 71 years; 67 females, 58 males) was used to illustrate the distribution of each of the TMS-EEG metrics in individuals without chronic pain and define the normal ranges. These TMS-EEG data derive from previously published studies [27,29,30] as well as ongoing work currently in preparation. This included identifying key statistical parameters, such as the median value, as well as the 25th and 75th percentiles, which serve as reference points for comparison with individuals without chronic pain.

### rTMS Treatment

Magnetic stimulation was delivered using a transcranial magnetic stimulator (MagPro R30, MagVenture A/S, Farum, Denmark) using a figure-of-eight coil (Cool-B35). Patients were seated comfortably in a reclining chair and instructed to stay as relaxed as possible during each session. Each rTMS session involved 30 trains of 100 TMS pulses given over 10 seconds (10 Hz), with 20-second pauses between trains, totaling 3,000 pulses over 15 minutes [1,29,30]. Stimulation intensity was set at 90% of the rMT. If patients reported any sensation in the hand during treatment, the stimulation intensity was reduced until no sensation was no longer felt. rMT was checked before each rTMS session to ensure proper stimulation. As for the TMS-EEG assessment, the rTMS treatment was delivered to the hemisphere contralateral to the side of the most intense pain. If no dominant pain side was reported, stimulation was delivered over the left hemisphere. All adverse events and reasons for treatment discontinuation were recorded throughout the study period. During the rTMS treatment, patients were permitted to continue their concurrent medication and other treatments. Supplementary Table 1 presents the pharmacological treatments at baseline and the end of the intervention.

### Statistical analyses

All statistical analyses were performed using the Statistical Package for Social Sciences (SPSS, version 25; IBM, Chicago, IL). All outcomes are reported as means and standard deviations, unless otherwise specified. Statistical significance was set to maintain a two-sided 5% level (using multiplicity adjustment where appropriate) for comparisons. The normality of data distributions for each clinical and neurophysiological outcome was assessed in Responders and Non-responders using the Shapiro–Wilk test and visual inspection of histograms for extreme violations. Separate two-way mixed model analysis of variance (ANOVA) was used to compare the clinical outcomes, including pain at the time of the visit (primary clinical outcome), in the last 24 hours, over the last 7 days, quality of life, BPI (pain severity and pain interference), HADS, sleep quality, fatigue, and number of medications. The within-subject factor was Time, and the between-subject factor was Group. An interaction effect between Group and Time was included in the models. Bonferroni-corrected multiple pairwise comparisons, and corresponding adjusted 95% confidence intervals (CIs) and P values were generated and reported in the results. The Greenhouse–Geisser approach was used to correct for violations of sphericity, and effect sizes (partial eta-squared η²_partial_) were calculated. For the neurophysiological outcomes, depending on the results of the normality test, Student’s t-tests (for normally distributed data) or the Mann–Whitney U test (a non-parametric alternative) was used to compare the groups. In total, 8 comparisons were made: GMFP, LMFP, ERSP, and ITC in three frequency bands in the central stimulated cluster. Levene’s test of equality of variance was used, and Cohen’s was used to assess the effect size (Cohen’s d). In addition to group comparisons, neurophysiological values were compared to the reference data. For each measure, the 25^th^ and 75^th^ percentiles of the reference distribution were calculated to define the interquartile range. The proportion of participants in each group falling below the 25^th^ percentile was computed and compared using a Chi-square test of independence (2×2 contingency table) to determine whether Responders were significantly more likely than non-responders to fall outside the reference range.

Additionally, Pearson correlation analyses, or Spearman correlation analyses when distributions were non-normal, were conducted to investigate associations between the percentage of pain reduction and the four neurophysiological outcomes that showed differences. Finally, as an exploratory analysis, we performed logistic regression with k-fold cross-validation (CV) to test whether baseline TMS-EEG features predicted responder status (Responder = 1, Non-responder = 0), following prior modelling work on TMS and EEG pain biomarkers [9]. To keep dimensionality small relative to the number of training samples per fold (given N = 43), we set three predictors: (i) GMFP, included as a broad measure of cortical excitability; (ii) alpha ERSP in the stimulated cluster, chosen to capture complementary aspects of alpha-band activity (supported by prior evidence for alpha oscillations in pain processing [9,15]) and (iii) alpha ITC in the stimulated cluster. All preprocessing and modeling were embedded in a leakage-safe scikit-learn pipeline (median imputation -> z-scaling -> penalized logistic regression). We fit elastic-net logistic regression (saga; class_weight=balanced), with hyperparameters (C, l1_ratio) tuned by an inner stratified CV. Because class counts allowed balanced splits, we used 5-fold cross-validation. Performance was obtained via stratified nested CV with an outer 5-fold split repeated 20 times (100 outer evaluations total). For each run, after inner-CV tuning, we selected a probability cutoff to target 80% sensitivity (as we gave more weight to correctly identify Responders as opposed to Non-responders), then evaluated this fixed cutoff on the held-out outer fold. Primary metrics were ROC-AUC (discrimination) and PR-AUC (positive-class performance with a baseline equal to the responder prevalence), and sensitivity at the 0.80-operating point. Results were summarized across the 100 outer evaluations as mean ± SD.

## Results

### Patient characteristics

The demographic, clinical characteristics, and main diagnosis are reported in Tables 1 and 2. At the end of the rTMS treatment, 20 were Responders (47%), and 23 were Non-responders (53%). Responders and Non-responders were similar in age, height, and sex distribution. The proportions of patients with nociceptive, neuropathic, and nociplastic pain, as well as primary and secondary pain types (ICD-11 classification), were similar between groups. Pain duration did not differ between groups (Responders: 9□±□7 years; Non-responders: 12□±□10 years).

**Table 1.** Demographic and clinical characteristics (mean ± SD)

| Characteristic | Responders | Non-responders |
| --- | --- | --- |
| Number of patients | 20 | 23 |
| Age (years) | 53±12 | 52±13 |
| Height (cm) | 173±10 | 174±8 |
| Weight (kg) | 91±20 | 85±18 |
| Sex (n) | F= 12; M= 8 | F= 14; M = 9 |
| Primary pain (International Classification of Diseases 11<sup>th</sup>) (n) | 6 | 8 |
| Secondary pain (International Classification of Diseases 11<sup>th</sup>) (n) | 14 | 15 |
| Nociceptive pain (n) | 10 | 8 |
| Neuropathic pain (n) | 4 | 7 |
| Nociplastic pain (n) | 6 | 8 |
| Pain duration (years) | 9±7 | 12±10 |

**Table 2.** Main diagnosis.

| **Responders (N=20)** | **Non-responders (N=23)** |
| --- | --- |
| Shoulder arthropathy | Traumatic nerve lesion (ulnar nerve) |
| Post-traumatic foot Complex Regional Pain Syndrome type 2 | Chronic widespread pain |
| Post traumatic spondilopathy | Cervical traumatic radiculopathy |
| Cervical spondilopathy | Traumatic lumbar radiculopathy |
| Fibromyalgia | Knee oosteoarhrosis |
| Neck arthrosis | Lumbar spine degenerative spondyloarthropathy |
| Shoulder arthropathy | Fibromyalgia |
| Chronic primary abdominal pain syndrome | Fibromyalgia |
| Fibromyalgia | Inflammatory bowel disease-related arthropathy |
| Postoperative arthropathy | Diabetic polyneuropathy |
| Fibromyalgia | Diabetic polyneuropathy |
| Fibromyalgia | Fibromyalgia |
| Shoulder pain with arthropathy | Fibromyalgia |
| Fibromyalgia | Postoperative arthropathy |
| Chronic primary cervical pain | Fibromyalgia |
| Arthritis due to Sjogren's disease | Chronic primary cervical pain |
| Polyneuropathy | Fibromyalgia |
| Knee oosteoarhrosis | Arthropathy |
| Lumbar spinal stenosis with radiculopathy | Post-operative |
| Inflammatory knee arthritis | Osteoarthritis |
|  | Chronic widespread pain |
|  | Lumbar spinal stenosis with radiculopathy |
|  | Amputation - phantom limb pain |

### TMS intensities, compliance, and side effects

The intensities used for TMS–EEG and rTMS are reported in Table 3. There were no significant differences between Responders and Non-responders in the intensity during the TMS–EEG assessment, and between the first and last rTMS sessions (all p > 0.05). Among the Responders, 14 patients received left-hemisphere rTMS and 6 received right-hemisphere rTMS. Among Non-responders, 18 received left-hemisphere stimulation and 5 received right-hemisphere stimulation. Out of the 12 total rTMS sessions, the average number of attendances to the rTMS sessions was 11.6□±□0.6 (97%) among Responders and 11.9□±□0.2 (99%) among non-responders. All missing sessions were isolated single sessions (seven Responders and two Non-responders). The maximum number of missed rTMS sessions per patient was two.

**Table 3.** Stimulation intensities during TMS–EEG and rTMS sessions (first and last treatment sessions) in Responders and Non-responders. Values are expressed as mean ± standard deviation. TMS–EEG intensities are reported as a percentage of the resting motor threshold (rMT) of the hand muscle twitch.

|  | TMS- EEG (% of the hand rMT) | rTMS intensity first session | rTMS intensity last session |
| --- | --- | --- | --- |
| Responders | 56±10% (95±5% of hand rMT) | 43±7% | 42±7% |
| Non-responders | 51±6% (94±5% of hand rMT) | 39±5% | 40±7% |

In 18% of the rTMS sessions, side effects were reported: The main side effect was headache (14%), followed by “feeling dizzy” (2%). Other side effects, including “A trembling or numb sensation or the feeling of electricity in a part of the body”, “Twitching movements of an arm, leg or other body part”, and “Unusual smells, tastes, or emotions”, “Unexplained confusion, drowsiness or weakness”, and “Unusual experiences - out of body experiences, a feeling of being distracted, your body feels different” were together, present in 2% of rTMS sessions. 61% of the side effects were reported during the induction week, and 39% were reported during the maintenance. There were no major or serious side-effects.

### Primary clinical outcomes

Compared to the Non-responders, pairwise comparisons showed that pain VAS scores were lower in Responders at day 3 of the induction phase (P = 0.011; 95% CI [-32.1 -9.9]), week 2 (P = 0.029; 95% CI [-26.8 -5.6]), week 3 (P = 0.001; 95%CI [-28.9 -10.8]), and from week 5 to week 8 (all P < 0.001; week 5: 95% CI [-35.6 -16.2]; week 6: 95% CI [-35.6 -15.3]; week 7: 95% CI [-36.9 -16.2]; week 8: 95% CI [-47.7 -30.9]) of the maintenance phase. Figure 2 shows the primary clinical outcome, and the ANOVA on VAS pain intensity, rated immediately after each rTMS session, revealed a significant Time × Group interaction (F_1,41_ = 9.923; P < 0.001; η²_partial_ = 0.19).

**Figure 1:**
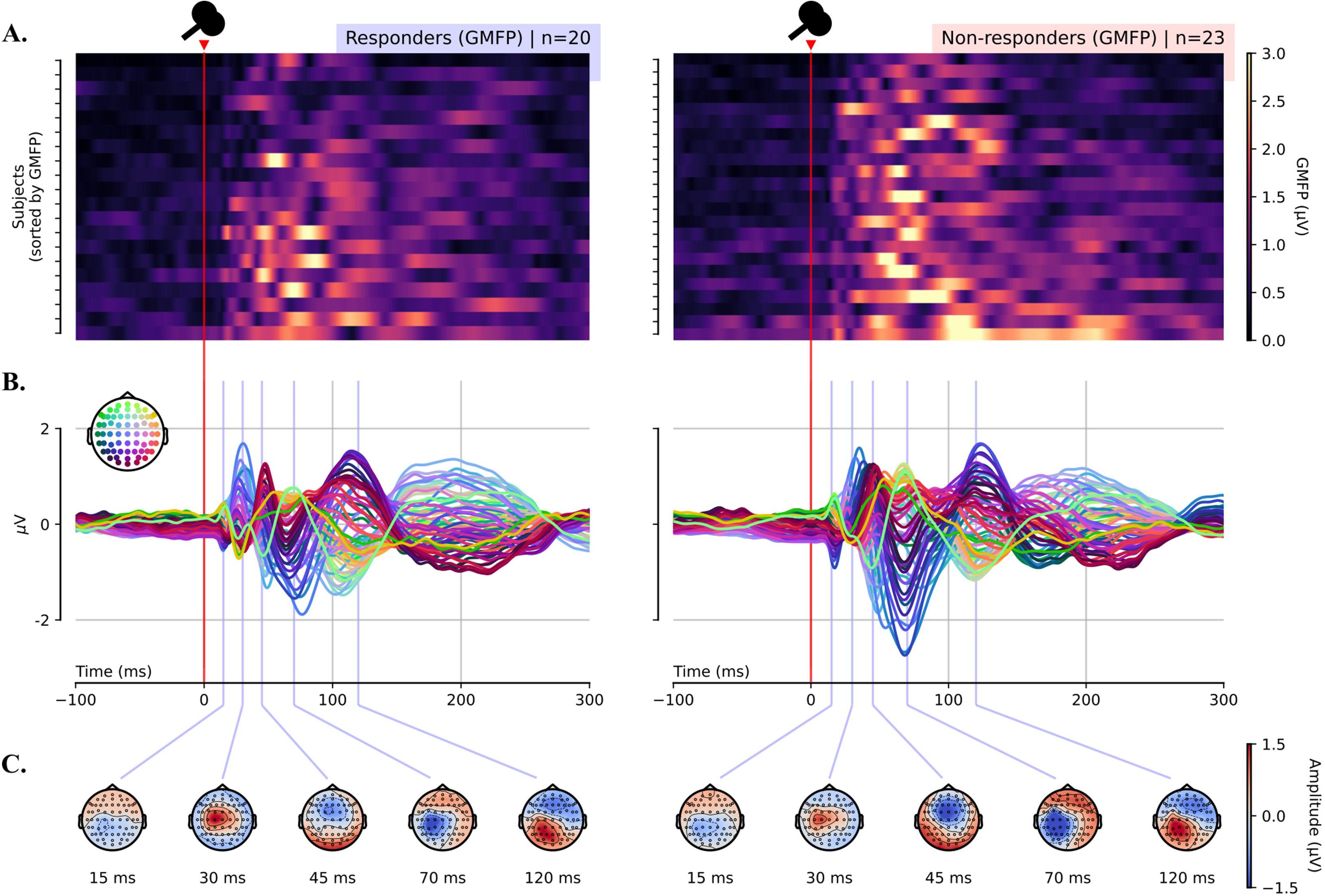
**(A)** Subject-level global mean field power (GMFP) after TMS for Responders (left) and Non-responders (right). Each row is one patient. Colour encodes GMFP amplitude (µV). **(B)** Butterfly plots of TEPs (one line per electrode) for Responders (left) and Non-responders (right). The red line indicates TMS onset (0 s). **(C)** Scalp voltage maps at 15, 30, 45, 70, and 120 ms post-TMS for Responders (left) and Non-responders (right). Warm/cool colours indicate positive/negative polarity (µV).

**Figure 2:**
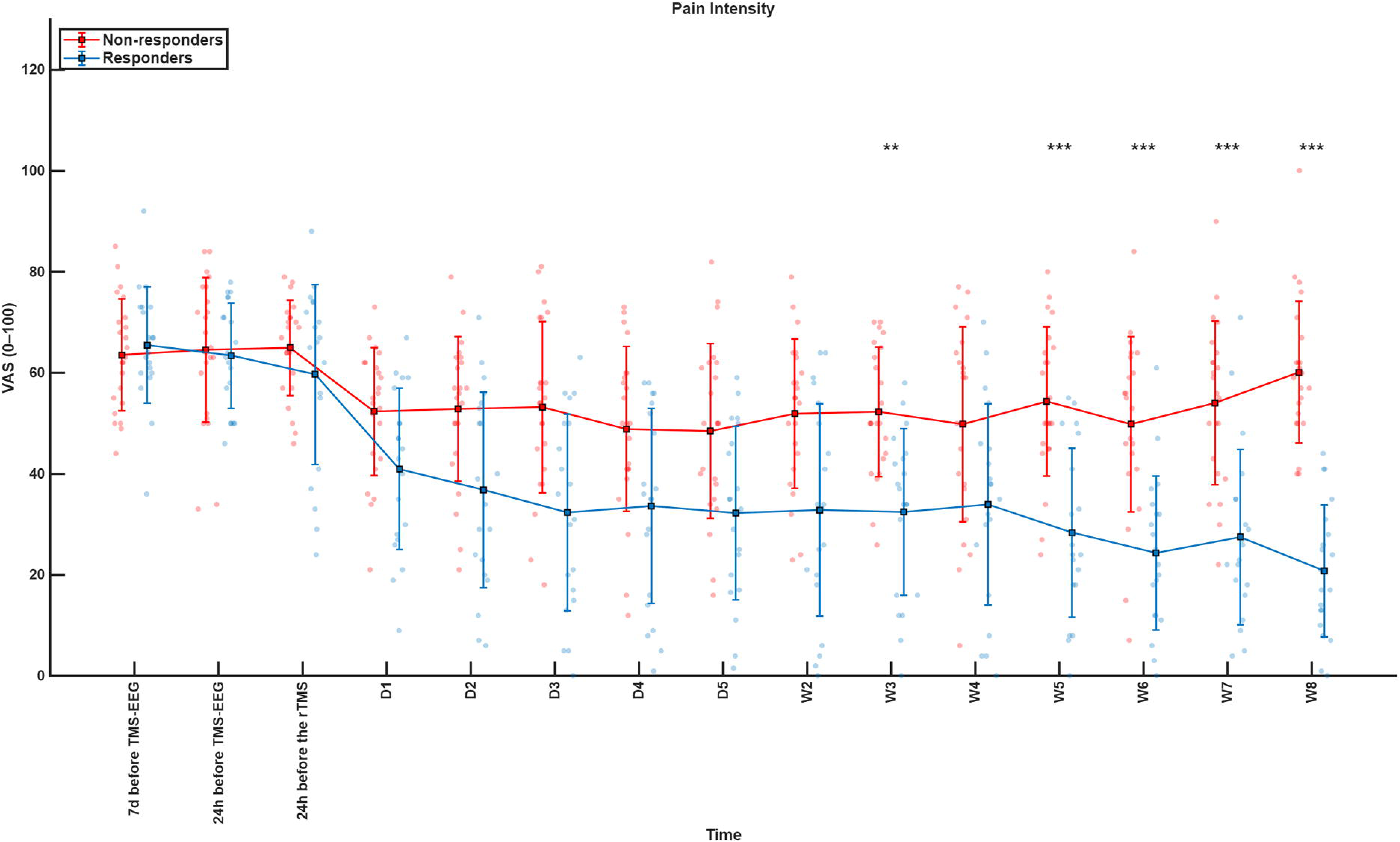
Pain intensity (Visual Analogue Scale, VAS: 0–100) across time points for Responders (blue) and Non-responders (red). Squares and error bars represent means ± standard deviations, with individual participant data shown as dots. The pre-treatment period includes assessments conducted 7 days and 24 hours before the TMS-EEG session, as well as 24 hours before the first rTMS session. The induction phase spans five consecutive treatment days (D1–D5), followed by a 7-week maintenance phase (W2–W8). Asterisks indicate statistically significant differences between groups at specific time points (*p < 0.05, **p < 0.01, ***p < 0.001).

### Secondary clinical outcomes

All secondary clinical outcomes are reported in Table 4 and Supplementary Figure 1. For the clinical outcomes assessed before and after the rTMS intervention, ANOVA revealed significant Time × Group interactions for average pain VAS intensity in the last 7 days (F_1,41_ = 20.464; P <0.001; η²_partial_ = 0.33), average pain VAS intensity in the last 24 hours (F_1,41_ = 8.663; P = 0.005; η²_partial_ = 0.17), pain severity (F_1,41_ = 18.271; P <0.001; η²_partial_ = 0.31), sleep quality (F_1,41_ = 7.517; P = 0.009; η²_partial_ = 0.16), fatigue (F_1,41_ = 5.961; P = 0.019; η²_partial_ = 0.13), and quality of life (F_1,41_ = 8.456; P = 0.006; η²_partial_ = 0.17). Post hoc pairwise comparisons revealed pain reduction in Responders compared with Non-responders over the last 7 days (P = 0.001; 95% CI [-31.4 -10.2] – Figure 3A), the last 24 hours (P = 0.001; 95% CI [-27.3 -8.2] – Figure 3B), pain severity (P = 0.001; 95% CI [-2.7 -0.9] – Figure 3C),), sleep quality (P = 0.013; 95% CI [4.1 32.5] – Figure 3D), fatigue (P = 0.011; 95% CI [-27.4 - 3.7] – Figure 3E), and an increase in the quality of life (P = 0.046; 95% CI [0.3 3.0] - Figure 3F).

**Figure 3:**
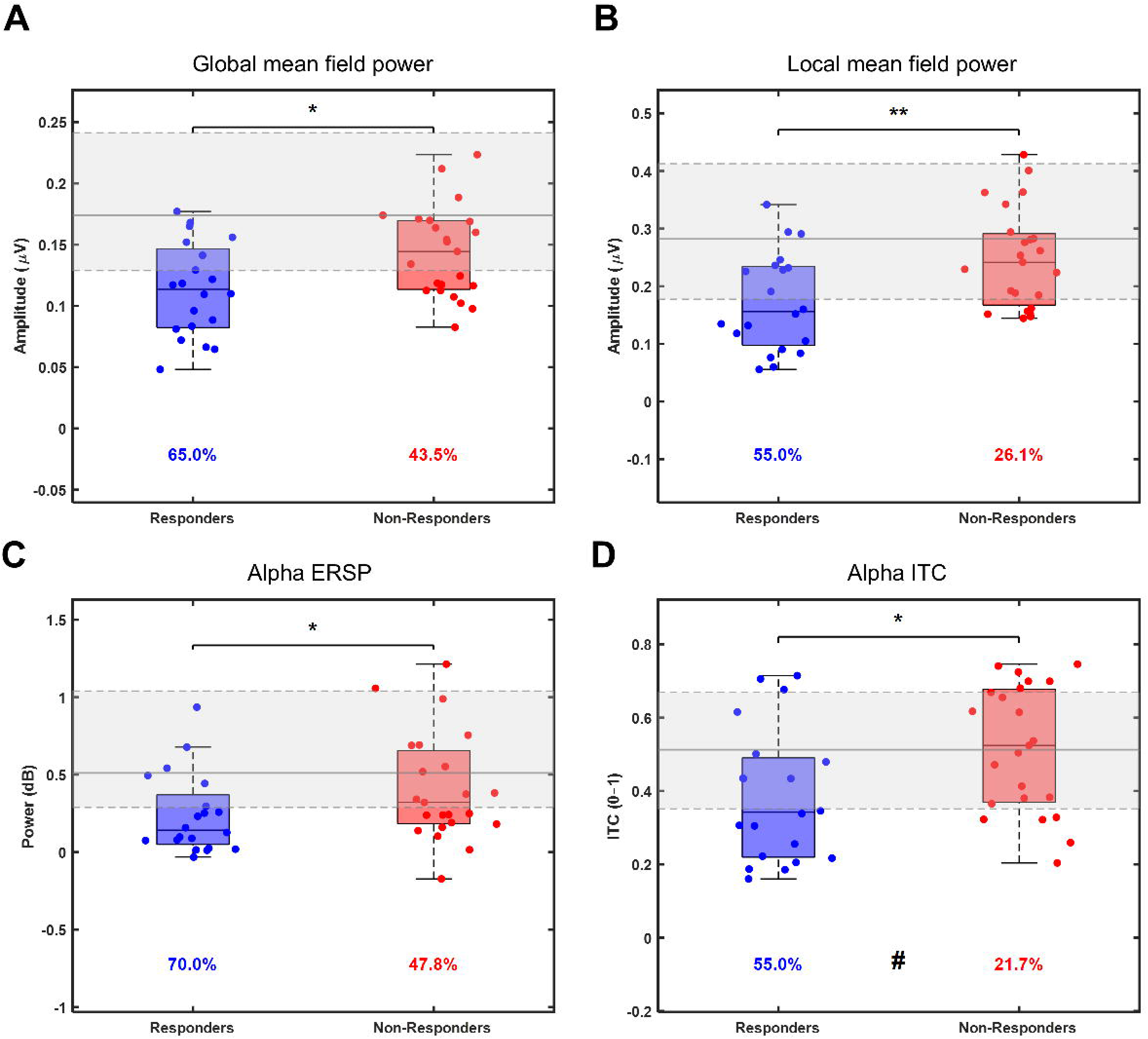
Neurophysiological outcomes in Responders (blue) and Non-responders (red) before rTMS treatment. Boxplots represent median ± quartiles, with individual participant data shown as dots. **A)** Global mean field power; **B)** Local mean field power; **C)** Event-related spectral perturbation (ERSP) in the alpha band; **D)** Inter-trial coherence (ITC) in the alpha band. Asterisks indicate statistically significant differences between groups (*p < 0.05, **p < 0.01).

**Table 4.** Mean (±SD) pain intensity in the last 24 hours, pain intensity in the last 7 days, number of painful areas, pain interference, pain severity, number of medications, quality of life, Hospital Anxiety and Depression Scale, sleep quality, and fatigue.

| Secondary clinical outcomes | Groups | Pre | Post |
| --- | --- | --- | --- |
| Visual analogue scale - Pain intensity last 24 h (0–100 mm) | Responders | 63±10 | 42±19*† |
| | Non-responders | 64±15 | 66±14* |
| Visual analogue scale - Pain intensity last 7 days (0–100 mm) | Responders | 66±12 | 44±19*† |
| | Non-responders | 63±12 | 65±16 |
| Number of painful areas (0–29) | Responders | 6.9±5.0 | 4.7±2.9 |
| | Non-responders | 7.2±5.3 | 6.2±4.4 |
| Pain interference (0–10) | Responders | 6.1±1.8 | 4.9±2.8 |
| | Non-responders | 5.6±1.6 | 5.5±1.5 |
| Pain severity (0–10) | Responders | 5.4±0.9 | 3.6±1.5*† |
| | Non-responders | 5.3±1.5 | 5.5±1.5 |
| Number of medications | Responders | 2.1±1.4 | 1.7±1.5 |
| | Non-responders | 2.2±1.0 | 1.9±1.2 |
| Quality of life EQ-5D (0–25) | Responders | 12.6±2.7 | 10.7±1.8*† |
| | Non-responders | 12.0±2.4 | 12.2±3.0 |
| Hospital Anxiety and Depression Scale (0–29) | Responders | 11.5±7.2 | 9.5±5.8 |
| | Non-responders | 13.8±6.0 | 13.1±6.2 |
| Visual analogue scale - Sleep quality (0–100 mm) | Responders | 49.4±26.0 | 68.2±20.1*† |
| | Non-responders | 53.6±31.2 | 50.9±28.9 |
| Visual analogue scale - Fatigue (0–100 mm) | Responders | 61.9±15.6 | 46.2±21.9*† |
| | Non-responders | 60.5±21.01 | 62.1±17.7 |

\* Significantly different from Pre within the group ( $P < 0.05$ ).
† Significantly different between groups ( $P < 0.05$ )

**Table 5.** Standardized coefficients and descriptive statistics of predictors used in the logistic regression model.

| Outcome | Standardized Coefficient | Cut-off at Other Variable Means (median) | IQR [25th–75th] |
| --- | --- | --- | --- |
| GMFP | -0.037 | 0.163 | [0.152, 0.176] |
| Alpha ERSP | -0.020 | 0.800 | [0.662, 0.975] |
| Alpha ITC | -0.036 | 0.673 | [0.571, 0.722] |

GMFP – global mean field power; ERSP – event-related spectral perturbation; ITC - inter-trial coherence

The number of medications used by patients did not significantly change during treatment in both groups (Time effect: F_1,41_□=□4.001, P□=□0.052, η^²^□□□□□□□□=□0.09; Time × Group interaction: F_1,41_□=□0.621, P□=□0.435, η^²^□□□□□□□□=□0.02; Supplementary Figure 2). The list of medications is provided in Supplementary Table 1.

For pain interference, the number of body areas with pain, and HADS, no differences were found (Supplementary Figure 2).

The PGIC scale revealed that, among Responders, 9 participants (39.1%) reported being “Much improved,” followed by 8 patients (34.8%) reporting “Minimally improved” at the end of treatment. However, 3 Responders (13.0%) reported “No improvement”. Non-responders most commonly reported “No improvement” (12 participants, 52.2%), followed by “Minimally improved” (7 participants, 30.4%). One Non-responder (4.3%) rated “Much improved”, while 2 (8.7%) as “Minimally worse”.

### TMS-EEG global and local mean field power

GMFP was lower in Responders compared to Non-responders (t_41_ = 2.571; P = 0.011; Cohen’s *d* = 0.786; Figure 3A). A total of 65% of Responders and 44% of Non-responders had values that fell below the 25^th^ percentile of the reference dataset (χ^²^ = 1.99; P = 0.158). Significantly lower LMFP was also identified in the Responder group in the stimulated cluster (t□□□=□2.858, P□=□0.007, Cohen’s *d*□=□0.874; Figure 3B). 55% of Responders and 26% of Non-responders had values below the 25^th^ percentile of the reference dataset (χ^²^ = 3.74; P = 0.053).

### TMS-EEG local oscillatory dynamics

In the electrode cluster centered around M1, alpha ERSP was lower in Responders compared with Non-responders (Mann-Whitney U-Test□=□2.070, P□=□0.038; Figure 3C) as well as alpha ITC (t□□□=□2.374, P□=□0.022, 95% CI [-0.24 -0.02], Cohen’s *d*□=□0.726; Figure 3D). For alpha ERSP, 70% of Responders and 48% of Non-responders had values below the 25^th^ percentile of the reference dataset (χ^²^ = 2.16; P = 0.142). For alpha ITC, 55% of Responders and 22% of Non-responders fell below the 25^th^ percentile (χ^²^= 5.07; P = 0.021).

### Correlation analysis

Low levels of reactivity and connectivity TMS-EEG metrics recorded pre-therapy correlated with post-treatment pain relief. The percentage of pain reduction significantly correlated with GMFP (Pearson r□=□0.40, P□=□0.008 - Figure 4A), LMFP (Pearson r□=□0.38, P**□**=□0.012 - Figure 4B), alpha ERSP (Pearson r□=□0.31, P□=□0.046 - Figure 4C) and alpha ITC (Spearman r□=□0.35, P□=□0.020 - Figure 4D).

**Figure 4:**
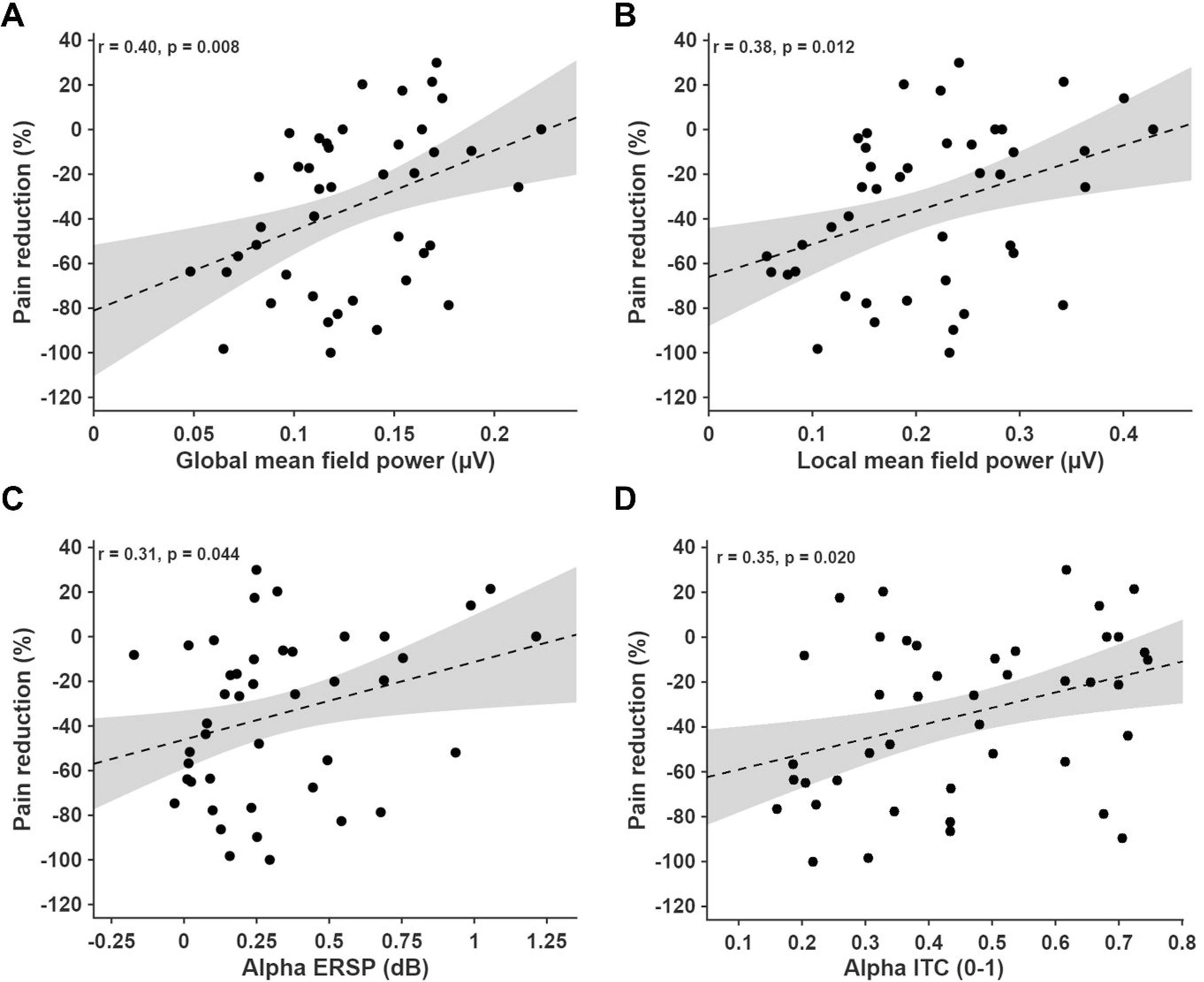
Correlation between percentage change in pain intensity and neurophysiological outcomes. Responders are shown in blue; Non-responders in red. **A)** Global mean field power (μV); **B)** Local mean field power (μV); **C)** Event-related spectral perturbation (ERSP) in the alpha band (dB). **F)** Inter-trial coherence (ITC) in the alpha band (0–1).

### Logistic regression for the prediction of response to rTMS

We combined three metrics of cortical reactivity, power-, and phase-based connectivity to explore the performance of a model to predict pain relief of M1 rTMS based on pre-therapy probing of M1. Using three predictors: GMFP, alpha ERSP, and alpha ITC, the model achieved acceptable performance across 100 folds. On average, ROC-AUC was 0.70 ± 0.18 and PR-AUC was 0.76 ± 0.15 (Figure 5). Table 4 summarises the standardised coefficients of the predictors, along with their median values and interquartile ranges. At the 80% cut-off, chosen to maximise sensitivity, sensitivity was 0.74 ± 0.26, with the expected drop in specificity (0.48 ± 0.28). These results suggest, on average, the model ranked a randomly chosen responder above a non-responder 74% of the time. Moreover, with Responders comprising ∼47% of the cohort, as the threshold for identifying Responders is varied, the model maintains ∼76% precision on average, versus a 47% baseline from random guessing, indicating acceptable positive-class performance. Lastly, using a cutoff chosen on the training split to target 80% sensitivity, the realised sensitivity on held-out data averaged 0.74 (∼3 out of 4 Responders correctly identified).

**Figure 5:**
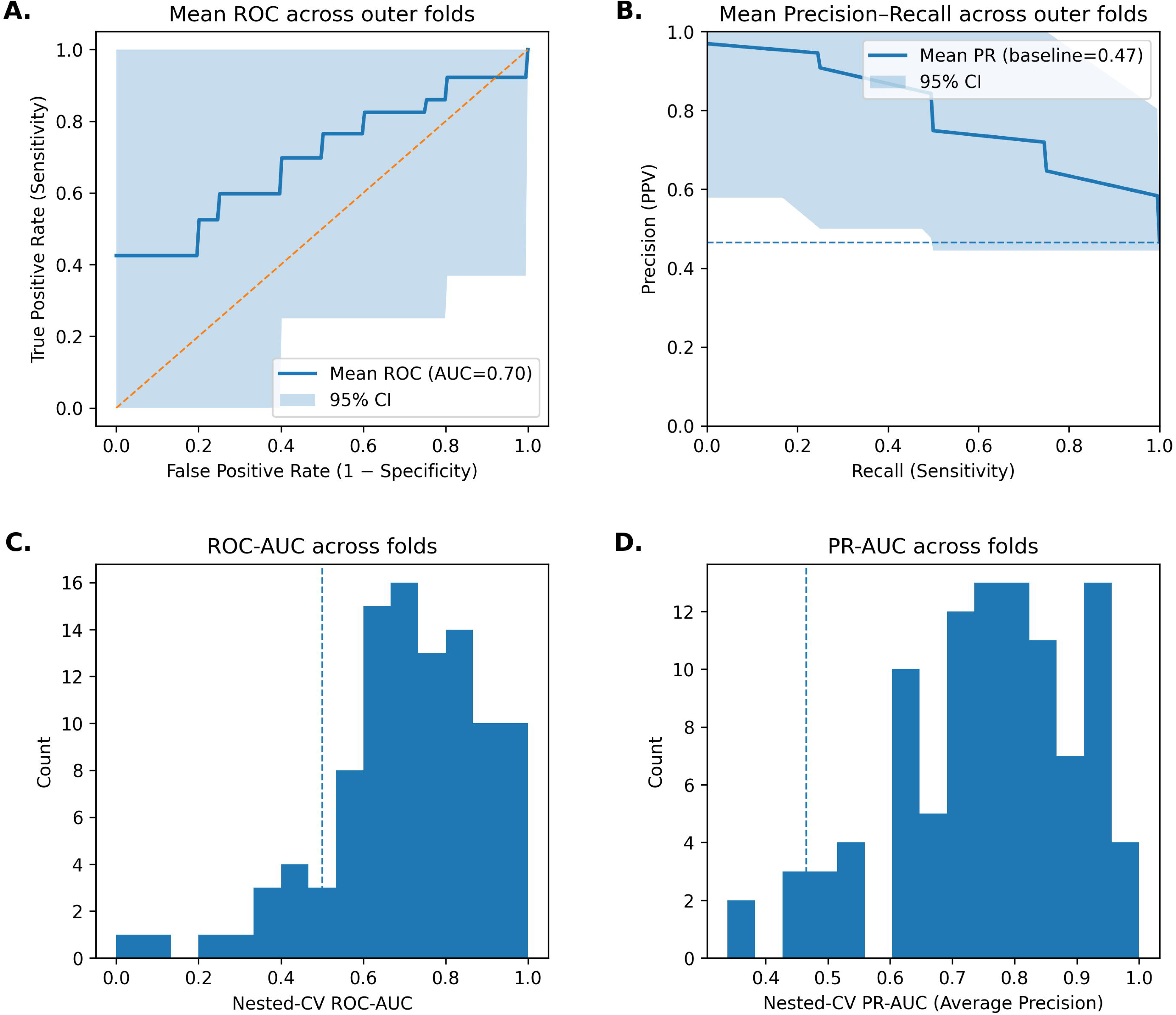
**A)** Mean receiver operating characteristic (ROC) curve with area under the curve (AUC = 0.70) and 95% confidence interval (CI). **B)** Mean precision–recall (PR) curve with average precision (PR = 0.47) and 95% CI; the dashed line indicates the baseline precision. **C)** Distribution of ROC-AUC values across 100 outer folds, with the mean indicated by the dashed line. **D)** Distribution of PR-AUC values across 100 outer folds, with the mean indicated by the dashed line.

## Discussion

The current study showed that patients responding to M1 rTMS treatment presented lower global and local cortical reactivity upon pre-therapy M1 probing. Furthermore, these patients showed reduced local TMS-evoked oscillatory dynamics in the alpha frequency compared to patients who did not respond to the rTMS treatment. These electrophysiological changes correlated with the percentage of pain reduction. Taken together, the findings suggest that lower pre-treatment global and local cortical reactivity and reduced alpha-band power and synchronization in the targeted region may reflect a neurophysiological state more prone to undergo adaptive neuroplastic changes, thereby increasing the likelihood of a favourable response to M1 rTMS.

### Clinical outcomes

In the current study, 47% of patients were responders to M1 rTMS. This percentage aligns with the recent findings of Quesada et al. [36], who reported a 47% response rate (17 out of 36 patients) and Hodaj et al. (27 out of 57 patients) [17], both using a ≥30% pain reduction criterion. Attal et al. identified 44% of patients as responders (22 out of 49) using a higher threshold of ≥ 50% pain reduction. Applying a ≥ 50% pain reduction threshold to our data, the responder rate was 39.5% (17 out of 43). Consistent with all previous studies [17,18,32,36], analgesic effects of rTMS mainly appeared after several days of rTMS treatment, indicating a cumulative therapeutic effect. In addition to reductions in pain intensity reported during the visit, the present study also found decreased pain severity, less pain in the previous 24 hours and at 1 week, and improved quality of life after rTMS. Conversely, no significant changes were observed in pain interference, the number of body areas affected by pain, depression, anxiety, or medication use. Furthermore, some patients reduced their own medication dosage during the trial, something previously reported in multicentre randomized sham-controlled trials on rTMS for chronic pain [44]. An important finding of the current study is that responders to M1 rTMS had various types of chronic pain conditions, not limited to neuropathic pain [1] and fibromyalgia [32]. Therefore, while most previous studies have focused on rTMS efficacy in specific chronic pain conditions [1,17,18,36], our results suggest that the analgesic effects of rTMS was not restricted to the specific pain diagnoses previously included in double-blind randomized trials.

### Global and local cortical reactivity

Consistent with the hypothesis that future responders to M1 stimulation had pre-treatment reductions in cortical reactivity, we found significantly reduced GMFP in responders compared with non-responders. These findings were confirmed by the presence of more GMFP results falling below reference ranges in the responder group. Cortical reactivity can be defined as the relationship between the strength of the stimulus and the magnitude of the evoked response, with GMFP representing the global amplitude of TMS-evoked brain activity across all electrodes [21,23]. The fact that responders also showed lower LMFP values around the stimulated M1 suggests that reduced local cortical reactivity may facilitate the therapeutic effects of rTMS. This may reflect a greater propensity for excitability enhancement through rTMS-induced plasticity, consistent with homeostatic mechanisms [2], which are more salient and likely to occur when baseline reactivity is low. Taking together, these findings suggest that reduced global and local M1 reactivity to single probing pulses of TMS may create a more favourable condition for upregulation through rTMS. In contrast, when global and local cortical reactivity is closer to reference levels, the capacity for further modulation may be limited, potentially resulting in weaker therapeutic effects.

### Local oscillatory dynamics

In the current study, patients responding to the rTMS treatment showed reduced TMS-evoked alpha power and phase coherence over the stimulated cortical area compared to non-responders. TMS-evoked oscillatory dynamics refer to the rhythmic activity generated by neuronal populations within a specific cortical region in response to the TMS stimuli, characterized by frequency-specific changes in power and phase from the pre-stimulus baseline [26]. These TMS-evoked metrics of connectivity provide insight into the functional state of the underlying cortical circuitry being probed [38]. The synchronization of the rhythmic activity across multiple frequency bands enables communication between spatially distributed brain regions through interareal phase locking [5]. This process creates time windows during which information can be integrated concurrently in sparse neuronal clusters, allowing complexity to emerge [14]. The observed efficacy of rTMS in chronic pain patients with the lowest alpha-band oscillatory dynamics may be interpreted in several ways. One possibility is that this pattern reflects larger disruptions in thalamocortical and corticocortical synchronization [42], potentially creating a neurophysiological environment more conducive to plastic reorganization by rTMS. Alpha-band oscillations are thought to propagate from higher-order to lower-order cortical regions and toward the thalamus via short-range supragranular feedback projections [16]. A reduction in alpha ERSP may indicate weakened local synchronization of neural circuits, while lower ITC suggests impaired phase alignment across repeated stimuli, indicative of reduced temporal precision and rhythmic coordination locally. Together, diminished alpha synchrony and temporal coordination may reflect a more "permissive" cortical state, facilitating greater susceptibility to modulation and plasticity through rTMS. Previous studies have shown that neuropathic pain and fibromyalgia are related to lower intracortical inhibition and facilitation [31], a sign of reduced GABA tonus within M1. rTMS to M1 has been shown to restore defective GABA-dependent intracortical inhibition coupled with pain improvement [32]. Alpha oscillations are implicated in large-scale cortical network integration, with phase locking and coherence modulating both local and long-range neural interactions [8,41].

Both connectivity metrics shown to be significantly low in Responders compared to Non-Responders to M1-rTMS were in the alpha band. An alternative explanation is that 10 Hz rTMS may be particularly effective in a specific subpopulation of patients who exhibit altered cortical oscillatory profiles near the alpha band frequency. Increasing evidence suggests that some individuals with chronic pain exhibit EEG frequency abnormalities, including reduced α- and low β-band activity (8–20 Hz), or increased high β-band activity (20–30 Hz) [35]. These frequency alterations are highly heterogeneous and highly variable across chronic pain patients [3]. A recent study reported that the closer an individual resting-state peak alpha frequency was to the rTMS frequency, higher were the changes of larger analgesic effects of 10Hz rTMS during tonic experimental pain [33], suggesting that rTMS may be more effective when delivered at or near an individual’s intrinsic M1 “natural” oscillatory frequency. In the current study, the observed reduction in alpha power and phase coherence among responders may reflect a cortical state closer to an optimal oscillatory dynamic, in which 10 Hz rTMS can more effectively induce plasticity and therapeutic effects. Conversely, it could be hypothesized that in non-responders, stimulation at different frequencies may be necessary to engage their intrinsic oscillatory dynamics and achieve comparable analgesic outcomes.

Another possibility concerns the target of rTMS. Individual differences in cortical pain processing may determine which cortical target, such as the M1, dorsolateral prefrontal cortex, or posterosuperior insula, is most responsive to neuromodulatory interventions. Experimental pain models in healthy participants have shown a polarization of pain-induced cortical suppression, with some individuals exhibiting reductions in excitability preferentially in one cortical area but not the other, reflecting marked inter-individual variability in pain-related cortical dynamics [28]. It is possible that, in the current study, the most appropriate cortical target for rTMS in non-responders would not be M1, but rather an alternative cortical area.

### Potential clinical applications

In an exploratory analysis, we used a supervised machine learning approach to test whether three baseline TMS-EEG features (ITC and ERSP Alpha in the stimulated cluster, and GMFP) could predict responder status to rTMS. We used a leakage-safe, stratified nested cross-validation framework with reduced dimensionality and penalized logistic regression to mitigate overfitting in a small sample size. Across 100 outer evaluations, discrimination and positive class performance of the model were acceptable on average, in which systematically lower ITC, ERSP, and GMFP predicted a higher likelihood of response to therapy. Moreover, with a cutoff chosen on the training split to target 80% sensitivity, the realized held-out sensitivity was acceptable, with the expected trade-off in specificity. These findings suggest that baseline TMS-EEG features may contain predictive signals for identifying rTMS responders, with an accuracy of about 75% (∼3 in 4 correctly identified), compared to the ∼45% response rate typically observed with the current use of M1 rTMS. In principle, an optimized model could support treatment selection by allowing clinicians to input patient-level values and estimate response probability, thereby informing individualized care. However, we do not yet claim to have such a model. Given the modest sample size and exploratory design (reduced dimensionality, penalized logistic regression, and stratified nested cross-validation), the results should be interpreted as indicating the feasibility of such approach, and viewed as a proof-of-concept. Larger, prospectively designed studies testing such an approach in clinical populations are needed before its clinical deployment is envisaged.

## Conclusion

This study provides the first evidence that baseline neurophysiological markers of global and local cortical reactivity and oscillatory dynamics recorded by probing the same target to be used for treatment are feasible and may predict the long-term response to rTMS in people with chronic pain. The presence of local decreased cortical responses may indicate a more maladaptive or dysfunctional network hub that is more likely to be positively influenced by neuromodulation. These findings support the potential utility of cortical reactivity and synchronization measures as a predictive strategy for identifying individuals most likely to benefit from rTMS interventions. Prospective trials with larger sample sizes specifically designed for this purpose are needed to test this new perspective and research paradigm.

## Data Availability

All data produced in the present study are available upon reasonable request to the authors

## Acknowledgements

The current study was supported by an ERC Horizon Europe Consolidator grant (PersoNINpain 101087925) and a Novo Nordisk grant (Grant NNF21OC0072828). The Center for Neuroplasticity and Pain is supported by the Danish National Research Foundation (DNRF121). Relevant to this study, Thomas Graven-Nielsen has received funding from the Lundbeck Foundation (R441-2023-232).

## Author contribution

**Enrico De Martino:** Conceptualization, Methodology, Formal analysis, Investigation, Data curation, Visualization, Writing - Original draft preparation; Project administration; **Margit Midtgaard Bach**: Investigation, Data curation, Writing - Review & Editing; **Anne Jakobsen**: Investigation, Writing - Review & Editing; **Stian Ingemann-Molden**: Investigation, Writing - Review & Editing; **Nahian S. Chowdhury**: Data curation, Writing - Review & Editing; **Bruno Andry Nascimento Couto**: Software, Formal analysis, Data Curation, Visualization, Writing - Review & Editing; **Adenauer Girardi Casali**: Software; Writing - Review & Editing; **Thomas Graven-Nielsen**: Writing - Review & Editing, Funding acquisition; **Daniel Ciampi de Andrade**: Conceptualization, Methodology, Visualization, Writing - Review & Editing; Project administration, Funding acquisition.

