## Supplemental material for "Lower pre-treatment TMS-evoked cortical reactivity and alpha-band oscillatory dynamics predict efficacy of primary motor cortex neuromodulation for chronic pain"

### **Supplementary material**

#### *Supplementary figure 1*

Secondary clinical measures assessed before and after rTMS treatment in Responder (blue) and Non-responder (red) groups. Bar plots display group means ± SD with individual data points overlaid. **(A)** Pain intensity in the last 24 h, **(B)** pain intensity in the last week, **(C)** pain severity, **(D)** pain interference, **(E)** number of body areas with pain, **(F)** number of daily medications, **(G)** sleep quality, **(H)** fatigue, **(I)** Hospital Anxiety and Depression Scale (HADS), and **(J)** EQ-5D quality-of-life score. Significant between-group differences are indicated (*p < 0.05, **p < 0.01, ***p < 0.001).

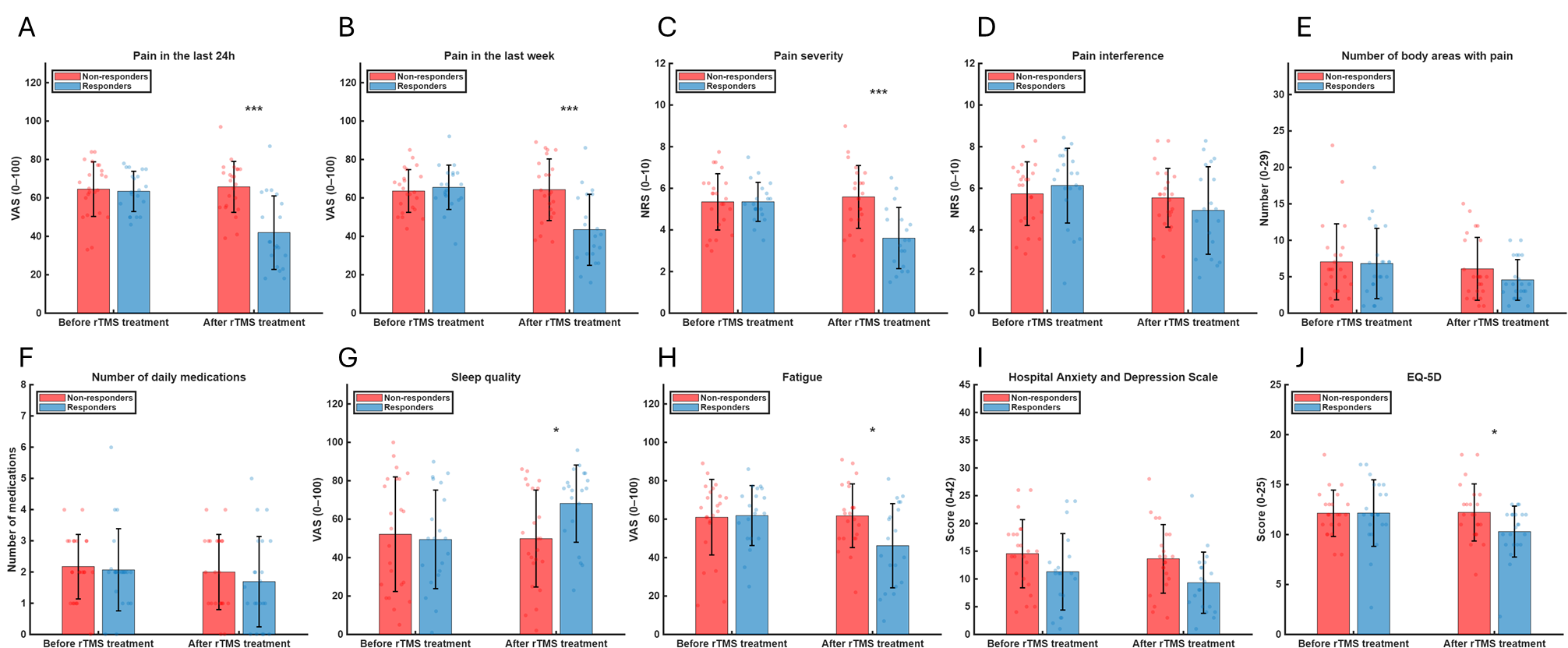

#### *Supplementary Table 1*

Table showing medication used in Responders (n=20) and Non-responders (n=23) before and after rTMS treatment.

| Medication | Group | Before rTMS | After rTMS |
| --- | --- | --- | --- |
| Acetaminophen | Responders | 17 | 15 |
|  | Non-responders | 14 | 9 |
| NSAIDs | Responders | 7 | 7 |
|  | Non-responders | 9 | 8 |
| Baclofen | Responders | 1 | 0 |
|  | Non-responders | 0 | 0 |
| Pregabalin | Responders | 4 | 4 |
|  | Non-responders | 1 | 1 |
| Gabapentin | Responders | 2 | 2 |
|  | Non-responders | 4 | 4 |
| Morphin | Responders | 2 | 2 |
|  | Non-responders | 2 | 2 |
| Tramadol | Responders | 4 | 3 |
|  | Non-responders | 4 | 2 |
| Codeine | Responders | 0 | 0 |
|  | Non-responders | 1 | 1 |
| Naloxone | Responders | 2 | 2 |
|  | Non-responders | 1 | 2 |
| Naltrexone | Responders | 4 | 4 |
|  | Non-responders | 2 | 2 |
| Amitriptyline | Responders | 2 | 3 |
|  | Non-responders | 0 | 1 |
| Nortriptyline | Responders | 0 | 0 |
|  | Non-responders | 1 | 1 |
| Duloxetine | Responders | 4 | 3 |
|  | Non-responders | 0 | 0 |
| Venlafaxine | Responders | 0 | 0 |
|  | Non-responders | 1 | 1 |
| Citalopram | Responders | 0 | 0 |
|  | Non-responders | 1 | 0 |
| Mirtazapine | Responders | 0 | 0 |
|  | Non-responders | 1 | 0 |
